# Who should get vaccinated first? An effective network information-driven priority vaccination strategy

**DOI:** 10.1101/2021.05.10.21256999

**Authors:** Dong Liu, Chi Kong Tse, Rosa H. M. Chan, Choujun Zhan

**Author notes:** Corresponding author Email address (Chi Kong Tse). This work is the results of a research project funded by City University of Hong Kong (Project Number 9229031).

## Abstract

Approval of emergency use of the Novel Coronavirus Disease 2019 (COVID-19) vaccines in many countries has brought hope to ending the COVID-19 pandemic sooner. Considering the limited vaccine supply in the early stage of COVID-19 vaccination programs in most countries, a highly relevant question to ask is: *who should get vaccinated first?* In this article we propose a network information-driven vaccination strategy where a small number of people in a network (population) are categorized, according to a few key network properties, into priority groups. Using a network-based SEIR model for simulating the pandemic progression, the network information-driven vaccination strategy is compared with a random vaccination strategy. Results for both large-scale synthesized networks and real social networks have demonstrated that the network information-driven vaccination strategy can significantly reduce the cumulative number of infected individuals and lead to a more rapid containment of the pandemic. The results provide insight for policymakers in designing an effective early-stage vaccination plan.

## 1. Introduction

The Novel Coronavirus Disease 2019 (COVID-19) pandemic markedly disrupts the normal daily life of people and severely impacts our economy. A variety of active intervention policies have been implemented to suppress the spreading of COVID-19, including social distancing [1], city lock-downs [2], contact tracing, quarantining [3], etc. The recent availability of effective vaccines has brought hope for a rapid recovery of our badly impacted economy and sooner resumption of normal life [4], despite the limited supply of vaccines in the foreseeable future presenting a major obstacle.

Vaccination is able to help our immune system actively produce antibodies which protect us against infectious diseases [5]. The use of vaccines has been proven effective in reducing the risk of being infected and saving lives during an epidemic outbreak. An effective vaccination strategy ought to immune people with a relatively high risk of getting infected so that the transmission of the virus can be halted as quickly as possible. For instance, once the “vital” individuals in a population are vaccinated and have gained immunity against the virus, the probability of a large-scale outbreak can be prevented due to the significantly reduced chance of spreading the virus to other individuals through these *relatively more important* individuals in the population [6]. To identify an effective vaccination strategy, some studies have adopted a network science perspective, combining a network model of the connected community and an epidemic model [7, 8, 9, 10], to provide a convenient framework to understand the impact of topological properties on the progression behavior of the epidemic. In particular, global and local network information can be incorporated in the derivation of effective vaccination strategies [11, 12, 13, 14, 15, 16, 17]. For global network information-based vaccination strategies, the network topologies are used by policymakers for developing vaccination strategies. For local network information-based methods, only partial network information is used. One of the basic assumptions made in much of the previous research on vaccination strategy is that vaccines have been approved for mass production and are provided adequately for large-scale vaccinations. However, despite the approval of vaccines for emergency use in the COVID-19 pandemic offering a defensive approach to protect vulnerable individuals [18], the vaccine supply is insufficient for now and in the foreseeable future. For instance, policymakers in Hong Kong are eager to pursue an effective vaccination strategy, and as of March 2021, the government has received the first batch of the vaccines including Sinovac^1^ and Comirnaty^2^ sufficient for covering around 20% of the entire 7.5 million population. Thus, determination of the priority groups for vaccination in the early stage of a large-scale vaccination is crucial to the effective utilization of the limited vaccine supply and to a rapid containment of the pandemic.

In this study, we use a network-based susceptible-exposed-infectious-recovered (SEIR) model to simulate the spreading dynamics of COVID-19 in a population. In identifying the optimal selection of priority groups to get vaccinated, we compare the network information-based targeted and random vaccination strategies in terms of their effectiveness in terminating the COVID-19 transmission. In particular, for a given network (representing a population), we choose one network property, namely node degree (defined as the number of connected neighbors of the node), to represent the relative extent of contacts of each node in the network. We introduce a feasible vaccination strategy based on the chosen network information. Furthermore, we use two intuitive indicators, namely the cumulative number of infected cases and the duration of progression, to validate the effectiveness of the vaccination strategy in comparison with the random vaccination strategy. Numerical results on both synthesized networks and large-scale real-world social networks show that vaccinating the groups consisting of nodes with higher degrees outperforms the random vaccination strategy in terminating the pandemic. Our results show that appropriately identifying priority groups based on their relative contact intensities, e.g., by professions, would lead to an effective vaccination strategy.

### 2. Methods

We first introduce the basic SIER model, and give a detailed description of two vaccination strategies to be compared.

#### 2.0.1. SEIR model and vaccination schemes

We adopt a network-based SEIR model [8] to describe the evolution and behavior of COVID-19 and contagion processes in a network. The SEIR model has been widely used in modeling dynamical behavior of pandemic and epidemic spreading [19, 20, 21]. Here, the network-based SEIR model differs from other models that describe the epidemic spreading dynamics by a set of differential equations [9] and generate the numbers of individuals in various states as functions of time. The nodes and the links in the network represent the individuals in a population and the contact frequency between two different individuals. In the SEIR model, a node in a network may assume one of the following four states:

- Susceptible (S): the node is susceptible to COVID-19 pathogen.
- Exposed (E): the node is infected through contact with infected nodes but cannot infect other susceptible individuals. Eventually, the exposed nodes become infected nodes.
- Infected (I): the node is infected and is able to infect susceptible individuals.
- Recovered (R): the node is recovered and gains full immunity.

The state transition is shown in Fig. 1. Initially, all the nodes in a network are in the susceptible state and once an infected node exists, the virus in the infected node starts to transmit to its neighboring nodes and begins to diffuse in the network. Let Ω_*i*_ be the set of neighboring nodes of node *i*. We denote *β*_*j*_(*t*) as the infectiousness value of node *j*, where node *j* belongs to Ω_*i*_ The probability that a susceptible node *i* becomes infected at time *t* as the sum of the infectiousness values of its neighbors, i.e.,

**Figure 1:**
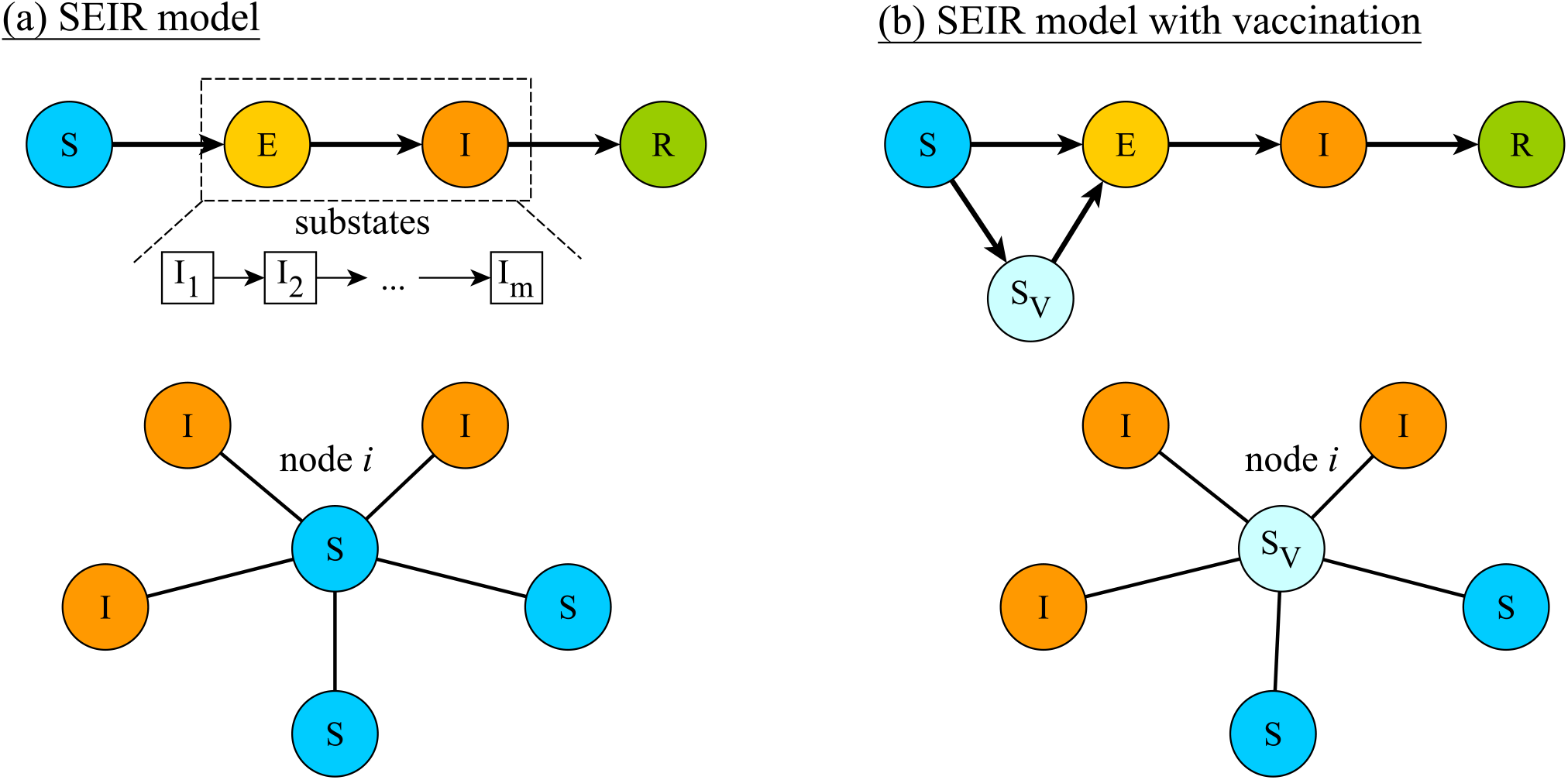
SEIR model without and with a vaccination program, as shown in (a) and (b), respectively.

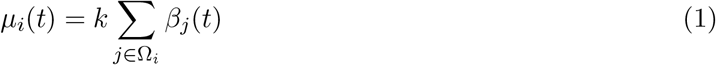

where *k* represents the relative infectiousness of the virus. Specifically, a larger *k* indicates that the spreading is more severe and has a relatively higher infection rate. To implement the infection process in the simulation, a random value between 0 and 1 is generated and compared with a parameter *µ*_*i*_(*t*). If the random value generated is smaller than *µ*_*i*_(*t*), the susceptible node enters the infected state. Furthermore, we also split the infected state of a node into *M* discrete substates, namely, *I*_*m*_ = *I*_1_, *I*_2_, …, *I*_*M*_, where various values of *β*_*j*_[*I*_*m*_] are given to characterize the infectiousness and determine the probability *β*_*j*_(*t*). Since an exposed node does not infect other nodes, the value of *µ*_*i*_(*t*) is set to zero in *M*_*v*_ (*M*_*v*_ *< M*) substates of the infected states. After node *j* recovers, it leaves the infected state and the value of *µ*_*i*_(*t*) turns to zero. Suppose node *j* is infected at time *t*_*i*_. Then, *β*_*j*_(*t*) can be written as a function of time *t*, i.e.,

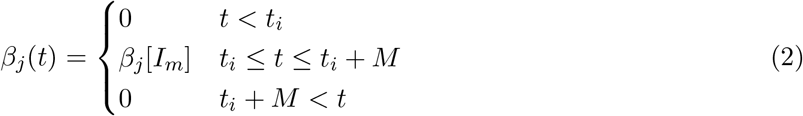

### 2.1. Vaccination strategies

To include the effect of vaccination in the model, we add one more state S_I_ to describe individuals who are immunized through vaccination. The main goal of vaccination is to protect individuals from being infected and thus to suppress the spread of the disease. Instead of simply removing the vaccinated nodes from the network [12], we introduce a new parameter called protection rate *r*_vacc_, which is linked to the effectiveness of the vaccination. Also, it is reasonable to assume that the protection effectiveness of the vaccine in the body would increase over time once vaccinated. Thus, the probability that a susceptible node *i* becomes infected at time *t* can be re-defined as

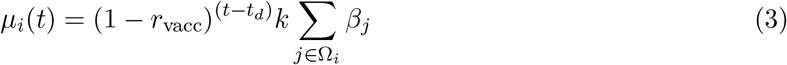

where the term 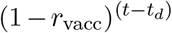 refers to the increased protection effect of the vaccine over time, and *t*_*d*_ is the time when the given node is vaccinated. In other words, as the node gradually gains immunity through vaccination, it is less likely to get infected by its neighboring infected nodes. It can be readily shown that the rate of increase of the percentage and peak value of infected nodes are significantly smaller in the presence of a vaccination program. In other words, vaccination significantly suppresses the spreading of the virus. (See Fig. S1 of Supplementary Appendix.)

Finding an effective vaccination strategy is of high relevance and significance in containing the pandemic, especially under the circumstance where the vaccine supply is limited in the early phase of a large-scale vaccination program and the disease is still aggressively propagating in the population. In order to validate the comparative advantage of a specific strategy of selecting priority groups for vaccination over a simple random selection strategy, we introduce two basic vaccination strategies, namely the network information-based vaccination strategy and the random vaccination strategy under the condition of limited provision of vaccine. Also, we assume a daily vaccination quota, denoted by *Q*_*V*_, to represent the maximum number of people getting vaccinated at each time interval.

The network information-driven vaccination strategy consists of two stages. At the beginning, the nodes in the network are ranked according to one given network property, such as node degree and node betweenness. Assuming the vaccine supply is enough for *N*_*V*_ individuals in the first stage of the simulation, the highest *N*_*V*_ ranked nodes are chosen as the priority group to get vaccinated. Then, in this stage, *Q*_*V*_ nodes are randomly selected from the priority group to get vaccinated at each time interval. Once the entire priority group has been vaccinated, the second stage begins, where vaccines are produced progressively. In the second stage, *Q*_*V*_ nodes are randomly selected from the rest of the unvaccinated nodes to get vaccinated at each time interval. Moreover, in the random vaccination strategy, no ranking procedure is performed and the vaccination is carried out by randomly choosing *Q*_*V*_ nodes at each time interval.

## 3. Results

Simulations for comparing the effectiveness of the network information-driven vaccination strategy with a random vaccination strategy are run on three types of synthesized networks, namely, BA scale-free networks [22], Watts-Strogatz (WS) small-world networks [23], and random networks. These networks contain 100000 nodes and the mean degree of the nodes in each network is fixed at 6. Furthermore, we use 12 realistic large-scale social networks to validate the effectiveness of the proposed network information-driven vaccination strategy for the early stage vaccination plan under limited vaccine supply. The parameters of SEIR-model with vaccination are set as follows. 1) *k* = 0.2 is chosen, corresponding to a mild level of severity of the pandemic spreading; 2) an 8-substage infection state (*M* = 8) is set based on real data fitting, where the infectiousness vector of an infected node is expressed as (*β*_*j*_[*I*_1_], *β*_*j*_[*I*_2_], *β*_*j*_[*I*_3_], *β*_*j*_[*I*_4_], *β*_*j*_[*I*_5_], *β*_*j*_[*I*_6_], *β*_*j*_[*I*_7_], *β*_*j*_[*I*_8_]) = (0, 0, 0.025, 0.075, 0.225, 0.25, 0.25). It can be seen that the first two substates correspond to exposed states, and the later substates having higher infectiousness value indicate stronger infectiousness; 3) 10 nodes in each test network are infected initially; 4) in the process of the vaccination, daily vaccination quota *Q*_*V*_ is set as 1% of the size of the test network; 5) a vaccination program is assumed to start after the disease has spread for a duration of delay time, denoted as *t*_*d*_. We set *t*_*d*_ = 30, and the vaccination strategy comes into the simulation model after *t* = *t*_*d*_.

### 3.1. Results on synthesized networks

For three test synthesized networks, we choose the highest *N*_*V*_ = *p*_*V*_ *N* ranked nodes as members of a priority group to be vaccinated, where *p*_*V*_ = 10%, 20%, 30% and *N* refers to the number of nodes in each network. In other words, the vaccine supply in the early stage can only cater for 10%, 20%, 30% of the population. Fig. 2 shows the growth curves of infected nodes in the three synthesized networks under different vaccination strategies. Specifically, these growth curves are obtained by averaging 2000 simulation runs. Compared with using random vaccination strategy, both the final value of the average infected population (i.e., cumulative number of infected individuals) and the average cessation time of the pandemic (when no more infected node is found) are smaller when the network information-driven vaccination strategy is adopted. Thus, the network information-driven vaccination strategy can lead to a more effective and rapid containment of the spread of the virus. Furthermore, applying the network information-driven vaccination strategy to BA scale-free networks yields the best performance among the three types of synthesized networks in terms of the distances among the growth curves, as shown in Fig. 2. The reason for such a finding is that compared with the other two types of networks, the BA scale-free network has a broader degree distribution. In a network with a broad degree distribution, more nodes with a high node degree (much larger than the mean degree) can be identified as members in the priority group to be vaccinated. The presence of these high-degree nodes would accelerate the transmission of the disease over the network and thus intensify the outbreak. Thus, vaccinating these nodes first has a significant effect on containing the spread.

**Figure 2:**
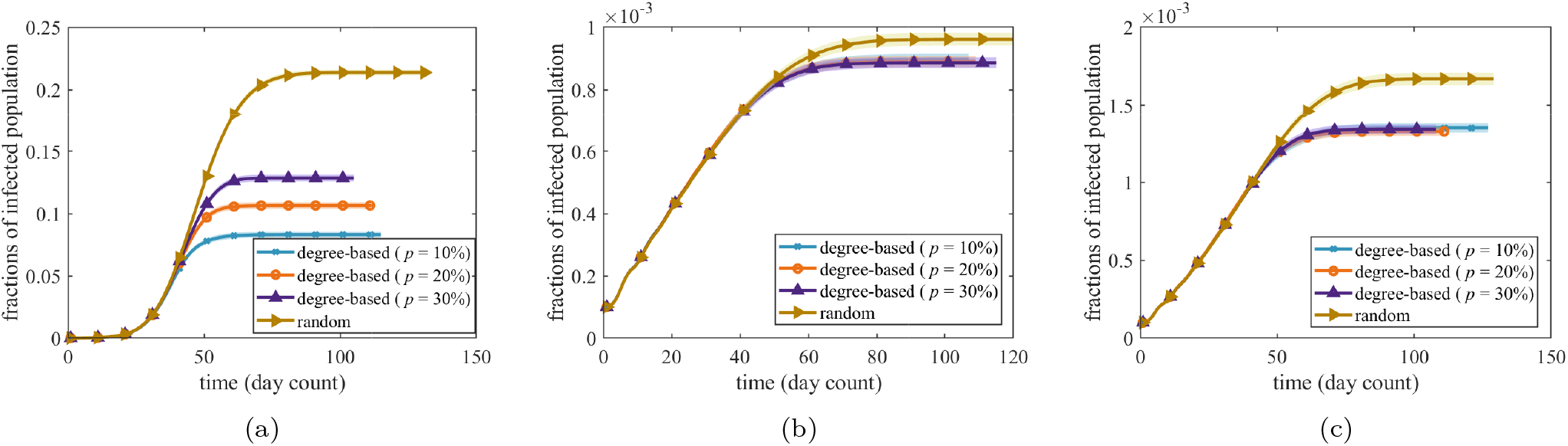
Comparison of growth of infected individuals in (a) BA scale-free network, (b) WS small-world network, and (c) random network using the network-information driven vaccination strategy and random strategy with 10%, 20%, 30% of the nodes in the test network chosen as the priority group to be vaccinated.

### 3.2. Results on real networks

We implement the proposed network information-driven vaccination strategy and the random vaccination strategy on 12 large-scale social networks. The network topologies are constructed from an online network dataset [24]. The number of nodes and mean node degree of these networks are in the range of 0.1 to 3 million, and from 4 to 16, respectively, which are similar to a real city-level population. For the network information-driven vaccination strategy, we choose the 20% highest node-degree nodes as members of the priority group to be vaccinated. Fig. 3 shows the growth curves of the infected populations for the two vaccination strategies in the real networks, which are obtained by averaging 2000 simulation runs. Comparing with the random vaccination strategy, the network information-driven vaccination strategy leads to reduced final sizes of infected population and shortened cessation time of the pandemic. Such results, consistent with the results on synthesized networks given in the last section, show the effectiveness of using network information for deriving a vaccination strategy for accelerating the containment of the pandemic.

**Figure 3:**
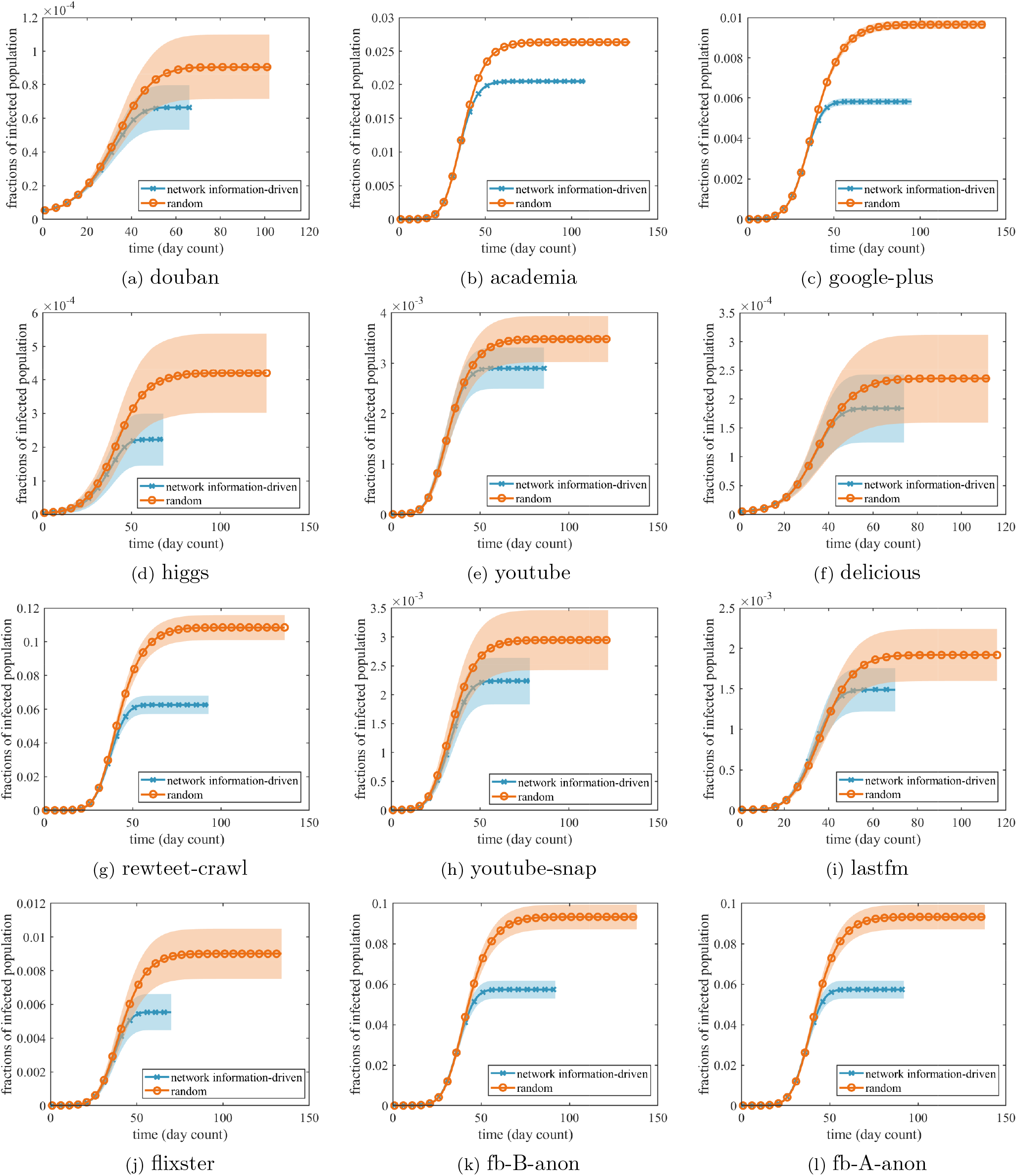
Comparison of growth of infected populations on 12 large-size real social networks using the proposed network-information driven vaccination strategy and random strategy with 20% of the nodes in the test network chosen as the priority group to be vaccinated.

To provide a quantitative comparison of the effectiveness of the network information based and random vaccination strategies, we propose two indicators, namely, the average cessation time *t*_end_ and the final value of the average number of infected nodes in the network *N*_*I*,final_. Here, *N*_*I*,final_ also represents the cumulative number of infected nodes. In Table 1, the first three columns show the basic network information. The fourth and fifth columns, and the seventh and eighth columns, give the *t*_end_ and *N*_*I*,final_ for the two vaccination strategies, respectively. Both *t*_end_ and *N*_*I*,final_ corresponding to the network information-driven vaccination strategy are larger than those corresponding to the random vaccination strategy. Furthermore, denoted as EFF, the *effectiveness* value(s) of the network information-driven vaccination strategy, defined as the percentage reduction in the size of the infected population and the duration of the pandemic in comparison with the random strategy, are given in the sixth and ninth columns in Table 1. Specifically, the cumulative infected population can be reduced by as much as 20% to 47% by adopting a network information based vaccination strategy, as given in the ninth column of Table 1.

**Table 1:** Average cessation time of the pandemic *t*_end_ and final value of the average number of infected nodes in the network *N*_*I*,final_ of 12 social networks. Subscript 1 denotes the network information-driven vaccination strategy where 20% highest node-degree nodes are chosen as the priority group for vaccination (*s*_1_), and subscript 2 denotes random vaccination strategy (*s*_2_). EFF is the percentage reduction of *t*_end_ and *N*_*I*,final_ using a network information-driven vaccination strategy compared with random strategy.

| Network | $N_{\text{node}}$ | $N_{\text{edge}}$ | $t_{\text{end},s_1}$ | $t_{\text{end},s_2}$ | EFF of $t_{\text{end}}$ | $N_{I,\text{final},s_1}$ | $N_{I,\text{final},s_2}$ | EFF of $N_{I,\text{final}}$ |
| --- | --- | --- | --- | --- | --- | --- | --- | --- |
| Douban | 154908 | 327162 | 44.31 | 50.34 | **11.98%** | 124 | 169 | **26.63%** |
| Academia | 200169 | 1398063 | 79.79 | 96.16 | **17.02%** | 38159 | 49118 | **22.31%** |
| Google-plus | 211187 | 1506896 | 65.94 | 102.61 | **35.74%** | 10844 | 17994 | **39.74%** |
| higgs | 456629 | 733647 | 53.74 | 79.12 | **32.08%** | 415 | 783 | **47.00%** |
| YouTube | 495957 | 1936748 | 55.83 | 72.40 | **22.89%** | 5407 | 6489 | **16.67%** |
| Delicious | 536108 | 1365961 | 44.82 | 53.41 | **16.08%** | 343 | 439 | **21.87%** |
| Rewteet-crawl | 1112702 | 2278852 | 46.36 | 53.60 | **13.51%** | 267 | 331 | **19.34%** |
| YouTube-snap | 1134890 | 2987624 | 55.66 | 72.7 | **23.44%** | 4170 | 5493 | **24.09%** |
| Lastfm | 1191805 | 4519330 | 50.49 | 64.53 | **21.76%** | 2779 | 3580 | **22.37%** |
| Flixster | 2523386 | 7918801 | 55.67 | 89.39 | **37.72%** | 10328 | 16791 | **38.49%** |
| Fb-B-anon | 2937612 | 20959854 | 64.75 | 100.66 | **35.6%** | 106852 | 173871 | **38.55%** |
| Fb-A-anon | 3097165 | 23667394 | 65.09 | 97.30 | **33.12%** | 116632 | 202228 | **42.33%** |

## 4. Discussion

Since early 2021, many countries have planned to launch vaccination programs of different scales according to the amount of vaccines available, and some have already launched an early phase of vaccination aiming to provide vaccines to certain priority groups in the population. While the selection of priority groups can be based on the need for protection, such as the elderly and medical workers, it is not always the most optimal strategy if the ultimate aim is to achieve an effective overall containment of the spread of the virus. In this study, we advocate a network information-driven vaccination strategy for expediting the containment of the pandemic when the vaccine supply is limited in the early phase of a vaccination program. Our findings show that for both large-scale synthesized and real social networks, a network information-driven vaccination strategy often achieves much smaller cumulative infected population and more rapid termination of the pandemic. Considering the severe impact of the COVID-19 pandemic on our life and economy, and the insufficient supply of vaccines in the foreseeable future, identification of appropriate priority groups for vaccination is critical to the effective and optimal utilization of the available vaccines. The key rationale behind the network information driven strategy is that individuals who have a relatively high contact intensity are more likely to get infected and are potential superspreaders. Then, by getting these individuals vaccinated first would provide significantly better protection to the entire population. Governments are therefore advised to consider setting priorities to different groups by profession that are ranked in the order of their relative contact intensities. Our findings show that the cumulative infected population can be reduced by as much as 20% to 47%, depending on the exact contact topology, by adopting a network information based vaccination strategy.

We have used a network-based SEIR model to simulate COVID-19 spreading in large-size social networks where the edge connecting two nodes represents the contact between these two nodes. The node degree is a natural parameter that can be used for identifying high-risk nodes as well as superspreaders of the virus in the network [25]. Once these nodes are protected by vaccination, the virus transmission chain would be significantly weakened. With limited vaccine supply, prioritizing the vaccination for the higher degree nodes has demonstrated clear advantage in contributing to the containment of the spread. Furthermore, as the degree distribution of any real-world population is never uniform or homogeneous, the percentage of population to be vaccinated in order to achieve herd immunity is expected to be significantly lower if a network information based vaccination strategy is adopted. In summary, our study shows that a network-based perspective will assist policymakers to develop proper early-stage vaccination strategies in a real population.

In practice, the node degrees of individuals and the degree distribution are not exactly known. Thus, to implement a network information-driven vaccination strategy, we may categorize the population into groups of different average degree values according to the respective professions and social circles. The node degree here can be interpreted as the number of contacts of an individual in a social network. We observe that employees in different work sectors have different levels of contact intensities. (See Fig. S2 of Supplementary Appendix.) In particular, workers in medical services, food services, accommodation, etc. have a higher risk of being infected as they contact more people in their work environment. However, vaccination strategies that give priority to elderly people [26, 27], e.g., early phase of Hong Kong’s vaccination program in March 2021, might not be as effective as a network information driven vaccination strategy. Recent statistical data shows that the elderly groups (60+ age groups) have much fewer social contacts compared with other age groups. (See Fig. S3 of Supplementary Appendix.) Thus, giving vaccination priority to the elderly groups is unlikely to result in more rapid containment of the pandemic.

In Hong Kong, for instance, around 23% of the population are employees in the medical sector, construction workers, import/export traders and wholesalers, hospitality and restaurant workers, and persons involved in transportation, postal and courier services [28]. Members of these groups undoubtedly have a relatively higher degree. According to the reported data, people in these groups were more likely to be infected [29]. Thus, the effectiveness of an early-stage vaccination program could be improved by implementing a network information-driven vaccination strategy such as the one proposed in this paper, i.e., identifying the priority groups to be vaccinated. It should be noted that our analysis was limited by the specific choice of social network for evaluating the effectiveness of the network information-driven vaccination strategy. Social networks are evolving dynamically and people in one social network might also exist in other social networks. In this study, effects of human migration among social networks are omitted. However, in the early stage of COVID-19 vaccination programs, coupling among different social networks is expected to be weak due to the imposition of travel restriction and social distancing. Thus, our findings are expected to be realistic.

## 5. Contributors

DL and CKT conceptualised the study and designed the model. All authors jointly designed and performed the simulations, analyzed the results, and wrote the paper. All authors revised the manuscript critically for important intellectual content, and approved the final version of the manuscript. All authors had full access to all the data in the study, and the corresponding author had final responsibility for the decision to submit for publication.

## Data Availability

The data used in this study is publicly available from the website shown below.

https://www.onetonline.org/find/descriptor/result/4.C.1.a.4?s=2\&a=1

## Supplementary Appendix

### Epidemic Progression in the Presence of Vaccination Program

To compare the epidemic progression in the absence and presence of a vaccination program, we randomly select 10 nodes to be infected initially and run simulations using the model for a 1000000-node Barabasi-Albert (BA) scale-free network [1] to obtain the numbers of individuals in different states at each time interval (day count). In particular, in the SEIR model with vaccination, we randomly choose 1% of nodes to be vaccinated in every time interval. Note that the choice of this percentage is immaterial as it only provides a convenient and consistent simulation parameter for comparison of different vaccination strategies. Fig. S1 shows the progression of the two epidemic models in the test networks, which are given by the percentages of nodes in the susceptible, infected (including exposed), and recovered states over time. The simulation ends when there is no newly infected node. According to Fig. S1, the percentage of the nodes in the susceptible state decreases slowly at the beginning and then drops dramatically to a fixed value. It is found that both the rate of decrement and the final percentage of susceptible nodes in the SIER model in the presence of a vaccination program are larger than those without considering vaccination. Moreover, the number of infected nodes escalates rapidly to a peak value and then reduces to zero at the end of the simulation while the number of recovered nodes keeps increasing in the entire propagation process. We also note that the rate of increase of the percentage and the peak value of infected nodes are smaller for the SEIR model with consideration of vaccination compared to the case in the absence of vaccination. It can be observed from this initial simulation run that vaccination significantly suppresses the spreading of the virus.

**Figure S1:**
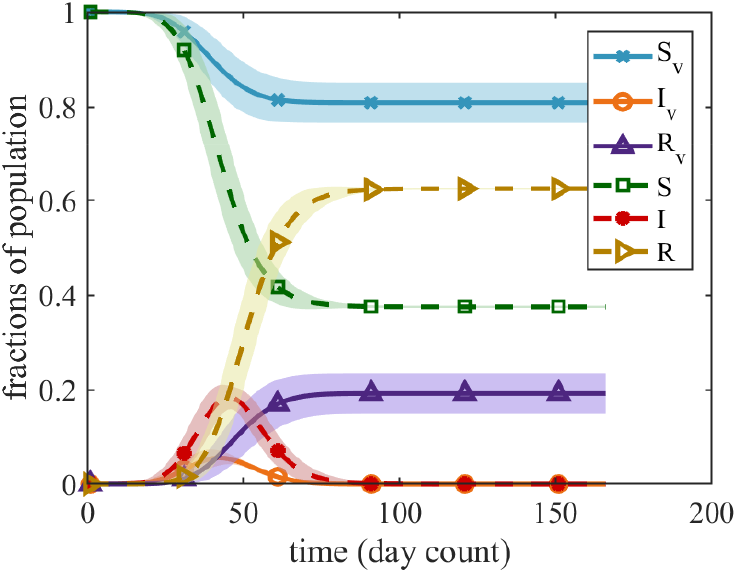
Comparison of progressions in the absence and presence of a vaccination program. “S”, “I”, “R” curves correspond to fractions of population in the susceptible, infected (including exposed), and recovered states in the absence of a vaccination program, and subscript “v” denotes progression in the presence of a vaccination program.

### Statistics of Contact Intensities

**Figure S2:**
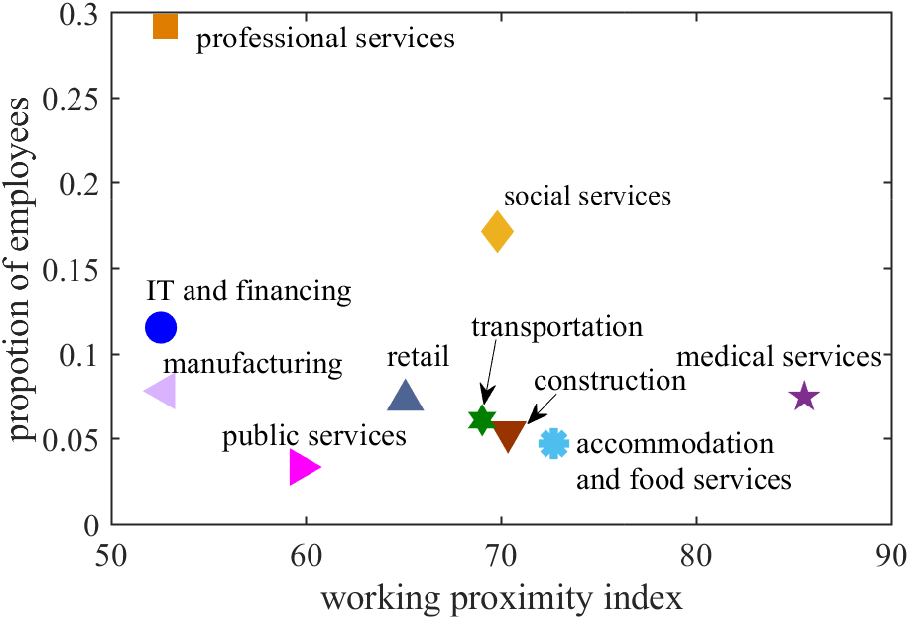
Proportion and working proximity index of employees in different work sectors. The working proximity index is calculated by averaging the scores given by the workers in answering the question of “how much does the job require the worker to be in contact with others” [2]. A larger working proximity index indicates a high contact intensity of workers in a particular sector.

**Figure S3:**
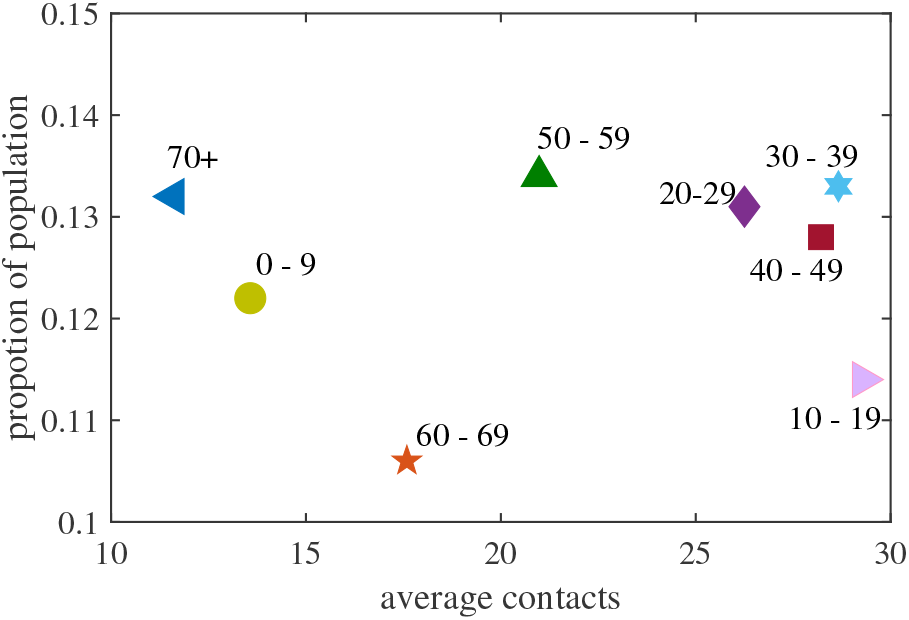
Proportion and average social contacts in different age groups. The data is from recent study [3], including BBC [4] and POLYMED datasets [5]. A large value of the average social contacts indicates a large number of contacts with other people.

## Footnotes

1 https://www.news.gov.hk/eng/2021/03/20210325/20210325_205321_468.html

2 https://www.info.gov.hk/gia/general/202102/27/P2021022700387.htm

